# Workplace infection control measures and perceived organizational support during the COVID-19 pandemic in Japan: a prospective cohort study

**DOI:** 10.1101/2022.06.06.22275988

**Authors:** Takahiro Mori, Tomohisa Nagata, Hajime Ando, Ayako Hino, Seiichiro Tateishi, Mayumi Tsuji, Ryutaro Matsugaki, Yoshihisa Fujino, Koji Mori, the CORoNaWork project

**Affiliations:** Department of Occupational Health Practice and Management, Institute of Industrial Ecological Sciences, University of Occupational and Environmental Health, Japan; Department of Work Systems and Health, Institute of Industrial Ecological Sciences, University of Occupational and Environmental Health, Japan; Department of Mental Health, Institute of Industrial Ecological Sciences, University of Occupational and Environmental Health, Japan; Disaster Occupational Health Center, Institute of Industrial Ecological Sciences, University of Occupational and Environmental Health, Japan; Department of Environmental Health, School of Medicine, University of Occupational and Environmental Health, Japan; Department of Public Health, School of Medicine, University of Occupational and Environmental Health, Japan; Department of Environmental Epidemiology, Institute of Industrial Ecological Sciences, University of Occupational and Environmental Health, Japan

**Keywords:** perceived organizational support, workplace infection control measures, health support, COVID-19, Japan

## Abstract

**Objective:** We investigated whether workplace infection control measures during the COVID-19 pandemic could increase perceived organizational support (POS).

**Methods:** This prospective cohort study was conducted in Japan from December 2020 to December 2021 using a questionnaire survey. There were 18,560 respondents at follow-up; we investigated 4,971 who rated low POS at baseline. The participants were asked a single question about POS and nine about workplace infection control measures. We determined the odds ratios (ORs) of high POS at follow-up using multilevel logistic regression analysis.

**Results:** The groups of 5–6 (OR=1.29; 95% confidence interval [CI], 1.05–1.57; *P*=0.014) and 7–9 workplace infection control measures (OR=1.54; 95% CI, 1.28–1.85; *P<*0.001) had significantly higher ORs than the group with 0–2 measures.

**Conclusions:** Health support for employees through workplace infection control measures can increase POS.

## Introduction

Defined as global beliefs concerning the extent to which an organization values employee contributions and cares about their well-being, perceived organizational support (POS) is an indicator of the relationship quality between employees and organizations.^1^ POS is known to be related to favorable outcomes for both. For example, high POS reportedly reduces job stress and burnout; it increases job satisfaction, work engagement, employee performance, organizational commitment, and organizational citizenship behavior.^2^ Even during the COVID-19 pandemic, studies have identified the association between POS and employee well-being-related indicators among health-care professionals and other workers. Research has reported that high POS relieves work stress, anxiety, depression, and burnout; it increases job satisfaction, organizational commitment, and work productivity.^3-10^ Increasing POS would, therefore, be of great significance to employees and organizations—even under the special circumstances of the pandemic.

POS has been found to be increased by several antecedent factors. According to a review by Sun, organizational factors (such as organizational fairness and justice), individual factors (such as employees’ traditional values and job status), and the relationship between organization and employees (such as values match between individuals and organization and leadership style) are reportedly antecedent factors for increasing POS.^2^ According to the definition of POS as the employee’s perception of organizational support for their well-being, providing workplace health support programs for employees may increase POS. In this regard, Grossmeier et al. investigated health and well-being best practices affecting POS among 812 organizations; they found that organizations with greater organizational and leadership support for employee health had higher POS.^11^ However, to our knowledge, no reports have shown that a workplace health support program for employees increased POS.

Under the COVID-19 pandemic, workplace infection control measures are intended to protect the health and life of employees, and so they may increase POS. It has been observed that adequate workplace infection control measures in Japan can cause positive changes at the individual employee level (such as positive effects on employee mental well-being and work performance and enhancing individual infection control measures).^12-14^ Considering these studies, we hypothesized that workplace infection control measures could improve both individual changes and POS, which reflects the relationship between organizations and employees.

If the results showed that POS increased as a result of workplace infection control measures, workplace health support programs for employees could increase POS. Workplace infection control measures during the COVID-19 pandemic could also lead to favorable outcomes for organizations and employees (such as improved work engagement and employee productivity) by increasing POS. Thus, we conducted a prospective cohort study to investigate whether workplace infection control measures could enhance POS.

## Methods

### Study design and participants

This prospective cohort study was conducted from December 2020 to December 2021 by a research group, the Collaborative Online Research on Novel-coronavirus and Work study (CORoNaWork study) of the University of Occupational and Environmental Health, Japan. Data were collected using a self-administered online questionnaire survey delivered via Internet survey company Cross Marketing Inc. (Tokyo, Japan). Details of the study protocol for the baseline survey have been previously reported.^15^ The participants included were workers aged 20–65 years at baseline; sampling was conducted by considering sex, occupation, and region of residence. In all, 33,087 were recruited; after excluding 6,051 who provided invalid responses, we included 27,036 participants in the baseline. We adopted the following criteria for invalidity: participants who completed the survey in extremely short response times; those who were shorter than 140 cm or weighed less than 30 kg; and those who gave inconsistent answers to multiple identical questions.

Those participants received a follow-up survey in December 2021, 1 year after baseline. In all, 18,560 participants responded to the follow-up survey (68.6% follow-up rate). The exclusion criteria for the present study were as follows: self-employed workers; workers in small or home offices; agriculture, forestry, and fishery workers; participants who retired or changed jobs after the baseline survey; and those who rated highly for POS in the baseline survey (our aim was to target participants with low POS at baseline). We also excluded participants who failed to respond to a question about POS at follow-up. We finally analyzed the data of 4,971 participants. Figure 1 is a flow chart for the study.

**Figure 1.**
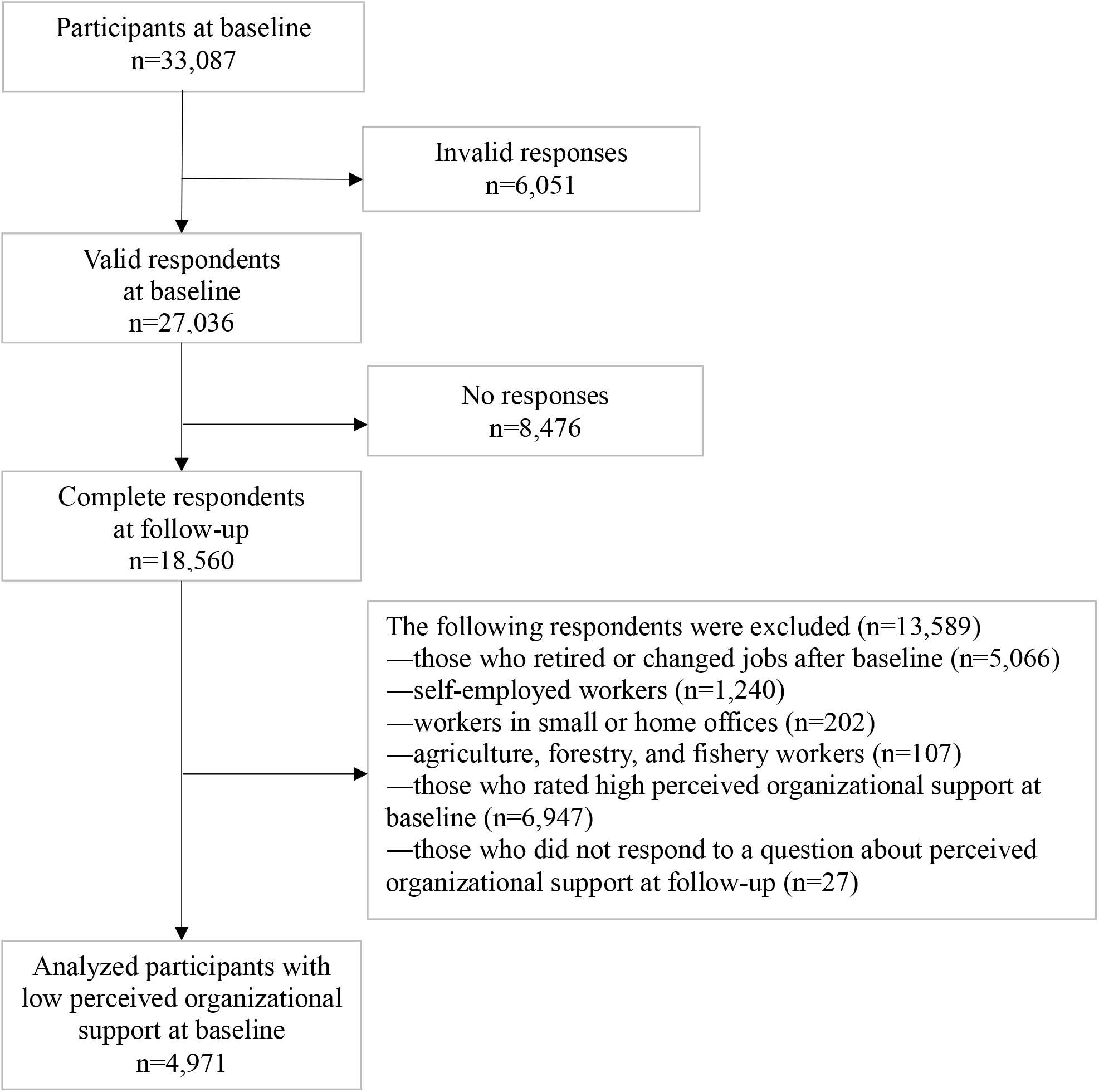
Flow chart for the study

This study was approved by the Ethics Committee of the University of Occupational and Environmental Health, Japan (Approval numbers: R2-079 and R3-006). Informed consent was obtained from all participants on the form of the website.

### Assessment of POS

We evaluated POS at baseline and follow-up with the following question, drawing on a previous study^16^: “Your company supports employees in finding a balance between active, productive working and healthy living.” Participants answered using a four-point scale (strongly agree, agree, disagree, and strongly disagree). We classified those who chose strongly agree and agree as having high POS and the others as low POS. As noted above, we selected participants with low POS ratings at baseline.

### Assessment of workplace infection control measures at baseline

With reference to a previous study, participants were asked at baseline to choose whether each of the following nine workplace infection control measures was implemented:^16^ prohibition or restriction of business trips; prohibition or restriction of visitors; prohibition of holding or limiting the number of people participating in social gatherings and meals; restriction on face-to-face meetings; requirement of always wearing masks during working hours; installation of partitions and change of workplace layout; recommendation for daily temperature checks; recommendation to telecommute; and requesting employees not go to the workplace when sick. We classified the number of workplace infection control measures into four categories: 0–2, 3–4, 5–6, and 7–9.

### Assessment of covariates

Covariates included demographics, occupation, job type, and number of workplace employees. Age was expressed as a continuous variable. We categorized annual equivalent income into four groups: *<*2.50 million, 2.50–3.74 million, 3.75–4.99 million, and ≥5.00 million yen (in 2021, US$1 was equivalent to 109.75 yen).^17^ Education was classified into three categories: up to junior high or high school, vocational school or college, and university or graduate school. We categorized occupation into 10 categories: general employee; manager; executive manager; public employee, faculty member or non-profit organization employee; temporary or contract employee; self-employed; small office/home office; agriculture, forestry, and fishing; professional occupation (e.g., lawyer, tax accountant, medical-related); and other occupation. In line with this study’s selection criteria, we excluded self-employed, small office/home office, and agriculture, forestry, and fishing. Thus, we classified occupation into seven categories. We categorized job type into three groups: mainly desk work; jobs mainly involving interpersonal communication; and mainly physical work. The number of employees in the workplace was classified into four categories: 1–9, 10–99, 100–999, and ≥1000 employees.

### Statistical analyses

We examined the association between the number of workplace infection control measures at baseline and high POS at follow-up. We determined age-sex adjusted odds ratios (ORs) and multivariate adjusted ORs using multilevel logistic regression analysis nested in the prefecture of residence to consider regional differences in the infection status of COVID-19. The multivariate model was adjusted for age, sex, income, education, occupation, job type, and number of workplace employees. We also conducted a trend test with the categories of the number of workplace infection control measures as continuous variables. A *P* value of less than 0.05 was considered statistically significant. We undertook all analyses using Stata Statistical Software (release 16; StataCorp LLC, College Station, TX, USA).

## Results

Table 1 presents the characteristics of our participants according to the number of workplace infection control measures. The group with greater workplace infection control measures had higher income and education, and it had more employees than the group with the least such measures. The higher the number of workplace infection control measures, the greater was the proportion of high POS at follow-up.

**Table 1.** Characteristics of participants at baseline according to number of workplace infection control measures

|  | Number of workplace infection control measures |  |  |  |
| --- | --- | --- | --- | --- |
|  | 0–2<br>n=1,265 | 3–4<br>n=884 | 5–6<br>n=1,070 | 7–9<br>n=1,752 |
| Age, mean (SD) | 47.2 (9.2) | 47.5 (9.2) | 47.2 (9.5) | 46.9 (9.3) |
| Sex, men | 727 (57.5%) | 486 (55.0%) | 587 (54.9%) | 965 (55.1%) |
| Annual equivalent income (million yen) |  |  |  |  |
| <2.50 | 368 (29.1%) | 199 (22.5%) | 175 (16.4%) | 255 (14.6%) |
| 2.50–3.74 | 396 (31.3%) | 279 (31.6%) | 275 (25.7%) | 435 (24.8%) |
| 3.75–4.99 | 297 (23.5%) | 219 (24.8%) | 332 (31.0%) | 542 (30.9%) |
| ≥5.00 | 204 (16.1%) | 187 (21.2%) | 288 (26.9%) | 520 (29.7%) |
| Education |  |  |  |  |
| Junior high or high school | 472 (37.3%) | 279 (31.6%) | 286 (26.7%) | 392 (22.4%) |
| Vocational school or college | 305 (24.1%) | 220 (24.9%) | 238 (22.2%) | 404 (23.1%) |
| University or graduate school | 488 (38.6%) | 385 (43.6%) | 546 (51.0%) | 956 (54.6%) |
| Occupation |  |  |  |  |
| General employee | 847 (67.0%) | 474 (53.6%) | 575 (53.7%) | 886 (50.6%) |
| Manager | 74 (5.8%) | 78 (8.8%) | 108 (10.1%) | 241 (13.8%) |
| Executive manager | 36 (2.8%) | 26 (2.9%) | 12 (1.1%) | 23 (1.3%) |
| Public employee, faculty member, or non-profit organization employee | 80 (6.3%) | 107 (12.1%) | 184 (17.2%) | 226 (12.9%) |
| Temporary or contract employee | 96 (7.6%) | 132 (14.9%) | 103 (9.6%) | 170 (9.7%) |
| Professional occupation (e.g., lawyer, tax accountant, medical-related) | 62 (4.9%) | 46 (5.2%) | 67 (6.3%) | 181 (10.3%) |
| Other | 70 (5.5%) | 21 (2.4%) | 21 (2.0%) | 25 (1.4%) |
| Job type |  |  |  |  |
| Mainly desk work | 637 (50.4%) | 413 (46.7%) | 524 (49.0%) | 957 (54.6%) |
| Mainly involving interpersonal communication | 216 (17.1%) | 219 (24.8%) | 290 (27.1%) | 367 (20.9%) |
| Mainly physical work | 412 (32.6%) | 252 (28.5%) | 256 (23.9%) | 428 (24.4%) |
| Number of workplace employees |  |  |  |  |
| 1–9 | 425 (33.6%) | 137 (15.5%) | 72 (6.7%) | 90 (5.1%) |
| 10–99 | 516 (40.8%) | 355 (40.2%) | 348 (32.5%) | 329 (18.8%) |
| 100–999 | 206 (16.3%) | 233 (26.4%) | 346 (32.3%) | 658 (37.6%) |
| ≥1,000 | 118 (9.3%) | 159 (18.0%) | 304 (28.4%) | 675 (38.5%) |
| Perceived organizational support at follow-up |  |  |  |  |
| High | 293 (23.2%) | 208 (23.5%) | 319 (29.8%) | 605 (34.5%) |

SD, standard deviation

Table 2 shows the association between number of workplace infection control measures and high POS. In the age-sex adjusted model, the groups with 5–6 (OR=1.41; 95% confidence interval [CI], 1.17–1.70; *P<*0.001) and 7–9 infection control measures (OR=1.75; 95% CI, 1.48–2.06; *P<*0.001) had significantly higher ORs than the groups with the least infection control measures. In the multivariate model, the same results were evident: 5–6 (OR=1.29; 95% CI, 1.05–1.57; *P*=0.014) and 7–9 infection control measures (OR=1.54; 95% CI, 1.28–1.85; *P<*0.001). We also observed a linear relationship between the categories of the number of workplace infection control measures and POS (P for trend < 0.001).

**Table 2.** Association between number of workplace infection control measures and high perceived organizational <u>s</u>upport

| Number of workplace<br>infection control measures | Age-sex adjusted |  |  | Multivariate adjusted* |  |  |
| --- | --- | --- | --- | --- | --- | --- |
|  | OR | 95% CI | *P* value | OR | 95% CI | *P* value |
| 0–2 | Reference |  | <0.001† | Reference |  | <0.001† |
| 3–4 | 1.02 | 0.84–1.26 | 0.817 | 0.97 | 0.78–1.19 | 0.749 |
| 5–6 | 1.41 | 1.17–1.70 | <0.001 | 1.29 | 1.05–1.57 | 0.014 |
| 7–9 | 1.75 | 1.48–2.06 | < 0.001 | 1.54 | 1.28–1.85 | < 0.001 |

\*Adjusted for age, sex, income, education, occupation, job type, and number of workplace employees
† *P* for trend
OR, odds ratio; CI, confidence interval

## Discussion

We found that among our participants, who had low POS at baseline, more workplace infection control measures during the COVID-19 pandemic significantly increased the OR for high POS 1 year later. This result supports our hypothesis of enhanced POS by health support for employees through active workplace infection control measures.

As to why workplace infection control measures increased POS among our participants, we considered that the employees recognized the organization valued their health by promoting initiatives that met employee needs. When employees believe they are receiving positive treatment through an organization’s voluntary actions rather than from external pressure (such as through trade union negotiations and government regulations), employees evaluate the organization more highly.^2^ Various workplace infection control measures are recommended by the Japanese government and specialized institutions^18,19^; however, it is up to an organization to implement such measures. It has been shown that the degree of infection control measures depends on the size of a company, job type, and other factors.^20^ Workplace infection control measures met employee needs during the COVID-19 pandemic, and they could reduce employees’ risk of and anxiety about infection.^12,13^ Therefore, employees may have recognized that companies are actively promoting health support for them through workplace infection control measures, and then POS may have increased.

Studies on the association between POS and work-related well-being during the COVID-19 pandemic have found that high POS reduces employee work stress and burnout; it improves work engagement, job satisfaction, and employee performance.^3-10^ There are reports of sufficient workplace infection control measures having positive effects on mental well-being and employee work performance.^12,13^ High POS has reportedly led to employees implementing infection control measures and more readily accepting COVID-19 vaccination.^16^ Infection control measures at the workplace have led to stronger individual infection control measures among employees during the COVID-19 pandemic.^14^ Thus, our results suggest that POS may have mediated the association between workplace infection control measures and those outcomes (i.e., mental well-being and undertaking individual infection measures). The mediating role of POS in this regard demands further investigation.

We believe that our results have important implications for implementing workplace health support program—even in non-pandemic situations. Providing a health support program that meets employee needs, such as the infection control measures in this study, can increase POS. To enhance POS when providing such a program, it is necessary to grasp employee needs and to communicate with employees so that they recognize the purpose of health support for them.^21^ Then, increasing POS will have a positive impact on management aspects of an organization, such as improved work engagement, better employee productivity, and reduced employee turnover tendency.^2,22^ Thus, implementing a workplace health support program can lead to both enhanced employee health and good outcomes for the organization. This study has some limitations. First, we evaluated POS using a simple question, and the measurement validity of POS was untested. We applied the same indicators in a previous study^16^; however, further research is needed to validate rigorously the measures for POS. In addition, evaluation of POS was undertaken by self-reporting, so it is possible that reporting bias occurred. We did, however, explain to participants that the survey would be anonymized, so we would expect the impact of this factor to be small.

Second, the workplace infection control measures we evaluated were not exhaustive. However, after reviewing a checklist of the Japanese government and the guides of specialized institutions,^18,19^ we discussed and decided on measures that many companies could adopt. Thus, we believe that our evaluation reflected the current state of COVID-19 measures at each company. In addition, some organizations may have changed their workplace infection control measures during the follow-up period, and so our results could be an underestimation of the actual situation.

Third, we did not consider other factors that may have increased POS. Supportive human resource practices (such as providing career development opportunities, offering equitable rewards, and creating a fair environment) also enhance POS^2^; however, we were unable to evaluate the effects of those factors.

Fourth, the duration of the effect of POS improvement through infection controls is unclear. We believe it is desirable to provide new health support programs for employees so they understand that they are receiving health support from the company.

In conclusion, our results show that employee health support provided by an organization through workplace infection control measures can increase POS. High POS can lead to good outcomes for both employees and organizations; thus, workplace infection control measures may have benefits beyond infection control. Our results suggest that providing a health support program that meets employee needs is significant from the perspective of increasing POS—even in non-epidemic situations.

## Data Availability

Data not available due to ethical restrictions

## Acknowledgments

The current members of the CORoNaWork Project, in alphabetical order, are as follows: Dr. Akira Ogami, Dr. Ayako Hino, Dr. Hajime Ando, Dr. Hisashi Eguchi, Dr. Keiji Muramatsu, Dr. Koji Mori, Dr. Kosuke Mafune, Dr. Makoto Okawara, Dr. Mami Kuwamura, Dr. Mayumi Tsuji, Dr. Ryutaro Matsugaki, Dr. Seiichiro Tateishi, Dr. Shinya Matsuda, Dr. Tomohiro Ishimaru, and Dr. Tomohisa Nagata, Dr. Yoshihisa Fujino (present chairperson of the study group), and Dr. Yu Igarashi. All members are affiliated with the University of Occupational and Environmental Health, Japan.

